# Parsing Neurometabolic Signatures of Multiple Sclerosis with MRSI and cPCA

**DOI:** 10.64898/2026.02.13.26346248

**Authors:** Narendiran Raghu, Mohammad Abbasi, Zeinab Tashi, Cristian Zamora, Shundene Key, Catherine D. Chong, Simona Niklova, Yuxiang Zhou, Edward Ofori, Benjamin B. Bartelle

## Abstract

Magnetic Resonance Spectroscopy Imaging (MRSI) offers spatially-resolved, neurometabolic information, acquired non-invasively at whole-brain scales from human subjects. Analysis of MRSI however, is extremely challenging. The metabolic information is highly convolved, and sparsely distributed across millions of spatial-spectral datapoints, allowing for little direct human interpretation. Conversely, the overall low signal-to-noise with high-intensity artifacts can confound unsupervised machine learning approaches. These technical barriers have left much of the potential of MRSI unrealized.

We acquired MRSI data from 4 human subjects with a diagnosis of multiple sclerosis (MS), incorporating experimental design into an informed machine learning approach. MRSI acquisitions were registered to anatomical MRI to label 105k spectra from brain tissue and 162 spectra from white matter hyperintensities (WMHs), an imaging biomarker associated with MS lesions. Spectral labels were then used in contrastive principal component analysis (cPCA) to filter artifacts and background features in the MRSI data from lesion salient features and clustered into statistically significant states based on features that could be interpreted from the original data. Our approach renders MRSI data into testable representations of neurometabolism, enabling the method for fundamental and clinical research.

**Graphical Abstract:** Analysis workflow for neurometabolic profiling of MS lesions. MRSI and anatomical MRI is acquired and processed in parallel for spectral data and anatomical labels. Spectra are then labeled and separated into experimental vs background data for contrastive PCA. Spectra are clustered for similarity, further labeled, and projected onto a brain atlas for a neurometabolic view.

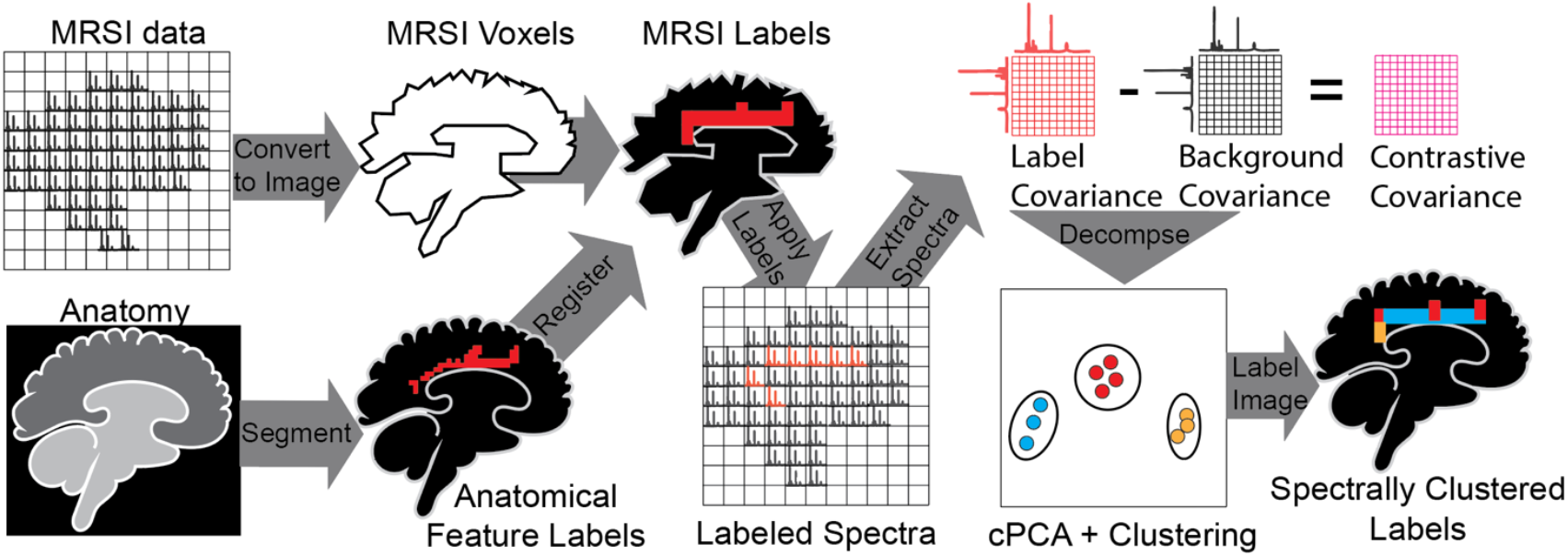

## INTRODUCTION

Neurometabolism is intertwined with neuroimmune function (1–3). Our evolving view of brain function now describes aspects of psychiatric disorders as neurometabolic imbalance, inflammation as a hypermetabolic state, and degeneration as energy collapse (4, 5). Some of the most significant biomarkers of inflammation TSPO, APOE and TREM2 are all, in fact, regulators of metabolism, specifically lipids (6, 7). While these proteins are resolved individually through targeted imaging, a change in their abundance marks a broader shift across hundreds of metabolites that can potentially be detected in a direct metabolic readout of the brain.

Metabolic profiling by fluid biopsy, mostly serum, has proven capable of classifying neurodegenerative disease and acute inflammatory states like multiple sclerosis (MS) using mass spectroscopy and nuclear magnetic resonance (NMR) (8, 9). In the case of MS, post-mortem analysis of lesions demonstrated distinct metabolic profiles compared to surrounding tissues and heterogeneity within the lesions themselves (10). This prior work establishes how assaying neurometabolism requires both molecular and spatial resolution, but acquiring such information from a living subject remains a frontier.

Magnetic resonance spectroscopy imaging (MRSI) combines the molecular resolution of NMR with the clinical imaging capacity of MRI, to deliver a spatial-metabolic readout of human subjects non-invasively and label-free. The method comes with substantial drawbacks, however. Brain tissue restricts the free tumbling of water and metabolites, reducing spectral resolution compared to NMR, at comparable field strengths. acquired from high-field clinical systems (3T+) under ideal conditions can only identify ∼12 of the most abundant metabolites with any confidence (11, 12). Despite the limitations, even single-voxel MRS has been used in clinical trials of MS, as the only non-invasive means of assaying lesions in human white matter (13).

While spectral peaks are degraded compared to liquid NMR, the tissue metabolome is still broadly represented in the MRS spectrum and enough to assay metabolic variation in aging and pathology, making MRS an increasingly valuable clinical method (14, 15). MRS spectral signatures can be leveraged to infer biology even if individual metabolites are not resolvable. Unbiased machine learning approaches based on dimensionality reduction and clustering, have been successful at using MRS to diagnose molecular pathologies, without quantifying metabolites (16). While machine learning approaches extract valuable clinical information from MRS interpretation remains a challenge and the methods require hundreds, if not thousands of samples to have predictive power (17).

Recent hardware advancements have allowed for parallel collection of whole-brain MRSI (12, 18). Here a single acquisition yields thousands of relevant spectra for analysis. This volume of data can only be analyzed computationally, and the tradeoff for rapid acquisition is low SNR with more artifacts. Like any NMR study, free water presents orders of magnitude more signal than metabolites and must be suppressed using destructive interference at a precise frequency based on the magnetic field. Anatomical structures like the ear canal and sinuses create localized field effects, resulting in severe spectral artifacts. Previous work has used machine learning approaches to filter informative signal from artifacts, including non-water-suppressed data using learned subspaces based on ideal MRS spectra (19, 20). This approach greatly improves SNR, at the cost of bias against metabolites not included in training data.

In this study, we sought to derive the maximum neurometabolic information from whole brain MRSI, to profile MS lesions from a small cohort of subjects and render this into human interpretable representations of metabolism. Previously, we utilized anatomy as “ground truth” to develop optimal representations of biology from spatial molecular data (21). Here we employ a similar strategy by collecting MRI data from each subject to resolve brain anatomy as tissue contrast and MS lesions as white matter hyperintensities (WMHs), then using this information to label MRSI spectra as lesion or undamaged brain tissues.

To incorporate all of this information into a feature learning analysis pipeline, we utilized contrastive principal component analysis (cPCA) (22, 23). Contrastive approaches leverage prior knowledge to highlight recurring “latent features” that are either unique to, or enriched within one dataset that are readily clustered or classified. Because cPCA is a linear dimensionality reduction method, the model is readily interpretable, producing eigenvectors similar to regular PCA. We contrast labeled WMH spectra from MS patients against non-lesion brain anatomy to generate several separable classes of spectra that could label anatomical scans or be projected onto a reference atlas to compare results across subjects in the same space.

## METHODS

4 subjects with a diagnosis of MS were scanned on the same 3Tesla clinical scanner using a 3D anatomical and MRSI sequence in a single session (Siemens 3T Prisma). MRI data was first collected for anatomical information and to highlight WMH’s using a T2 FLAIR sequence. MRSI was acquired using an echo planar imaging (EPI) sequence with broad-spectrum spatial-spectral (SPSP) excitation.

| Subject | Total Spectra | Brain Spectra | WMH Spectra |
| --- | --- | --- | --- |
| 1 | 131,072 | 20,216 | 78 |
| 2 | 131,072 | 27,280 | 57 |
| 3 | 131,072 | 27,720 | 21 |
| 4 | 131,072 | 30,659 | 6 |

### Data Acquisition, Preprocessing, and Registration

MRSI data of reference water peak data and water suppressed metabolite data, upon acquisition was converted into spatial information using a fast Fourier transform (FFT) prior to encoding in DICOM format. Upon completion of the study, these data were then extracted as 131072 × 512 matrices of free induction decay (FID) time domain data, which was converted into a spectrum of frequencies using the FFT function in NMRglue (24). For this analysis, only the real component of the transform was rebuilt into the original matrix dimensions used for acquisition. Spectral datasets were then converted into images for registration by summing the area under the curve into a single value image reconstructions, equivalent to low resolution proton weighted MRI.

In parallel, high-resolution MRI FLAIR scans were processed to identify WMH’s. Brain tissues and WMH’s were manually segmented from each anatomical scan and stored as labeled matrices. To align MRSI and MRI data spatially, the high-resolution anatomical images were downsampled to match the lower spatial resolution of the MRSI data. Image registration was performed using ANTsPy, a Python interface to the Advanced Normalization Tools framework, warping the FLAIR images onto the MRSI space (25). This transformation was then applied to the brain and WMH segmentation labels, enabling the identification of MRSI spectra corresponding to hyperintense lesions versus normal background tissue. MRSI data was then combined with MRI derived segmentation labels and the data was rearranged into a 2D matrix of 268M datapoints across 524,288 spectra, with 105,875 labeled brain spectra and 162 labeled WMH spectra.

### Feature Extraction and Contrastive Learning

All analysis was performed in Python using NumPy and Scikit-Learn. The number of components for PCA and cPCA were determined using an elbow plot for further clustering, however cPCA visualizations were performed using 2 components as recommended by the package. For cPCA, brain and WMH labeled MRSI spectra were parsed, using brain labeled tissue as the background while analyzing spectral variations in MS patients. To gain lesion-specific insights, a second cPCA was run, isolating hyperintense regions and comparing them against only the control subjects. The cPCA contrast hyperparameter (α), which controls the degree of contrast enhancement in cPCA, was systematically varied from 0 (equivalent to standard PCA) to 100 (high contrast emphasis). The optimal α value was determined based on how well different clusters were separated within the latent space.

### Clustering and Spatial Mapping

To further categorize spectral variations, K-means clustering was applied to the latent features derived from cPCA. The optimal number of clusters was determined based on inertia using an elbow plot analysis (Scikit-Learn), which identified three clusters as the most meaningful partitioning of the data. To project data onto a common framework, the anatomical images were registered to the ICBM 452 T1 Atlas space using ANTsPy, then the same forward transformation was applied to the cluster labels (26). Finally, the anatomical projections were visualized in both 2D and 3D using Napari, providing a visual representation of how spectral clusters align with known neuroanatomy (27).

## RESULTS

### MS lesions are readily detected in human subjects by MRI

To collect anatomical ground truth to inform MRSI analysis, we used T2-FLAIR MRI, a method with high SNR and sensitivity to MS lesions. In all subjects, white matter appeared as a hypointense region compared to other tissues, with MS lesions appearing as white matter hyperintensities (WMHs) (**Figure 1A**). These were largely proximal to the lateral ventricles and corpus callosum, with some in deeper brain regions. WMHs were manually segmented from MRI and co-registered to a brain atlas to compare lesions across multiple subjects (**Figure 1B**). Lesion patterning appeared initially random across the white matter, but subjects with high lesion burden had contiguous WMH pixels that could be interpreted as large lesions or clusters. In a comparison of lesion sizes and counts across subjects, variance increased with lesion burden, with Subject 1 having both the largest and smallest detectable WMHs (**Figure 1C**).

**Figure 1:**
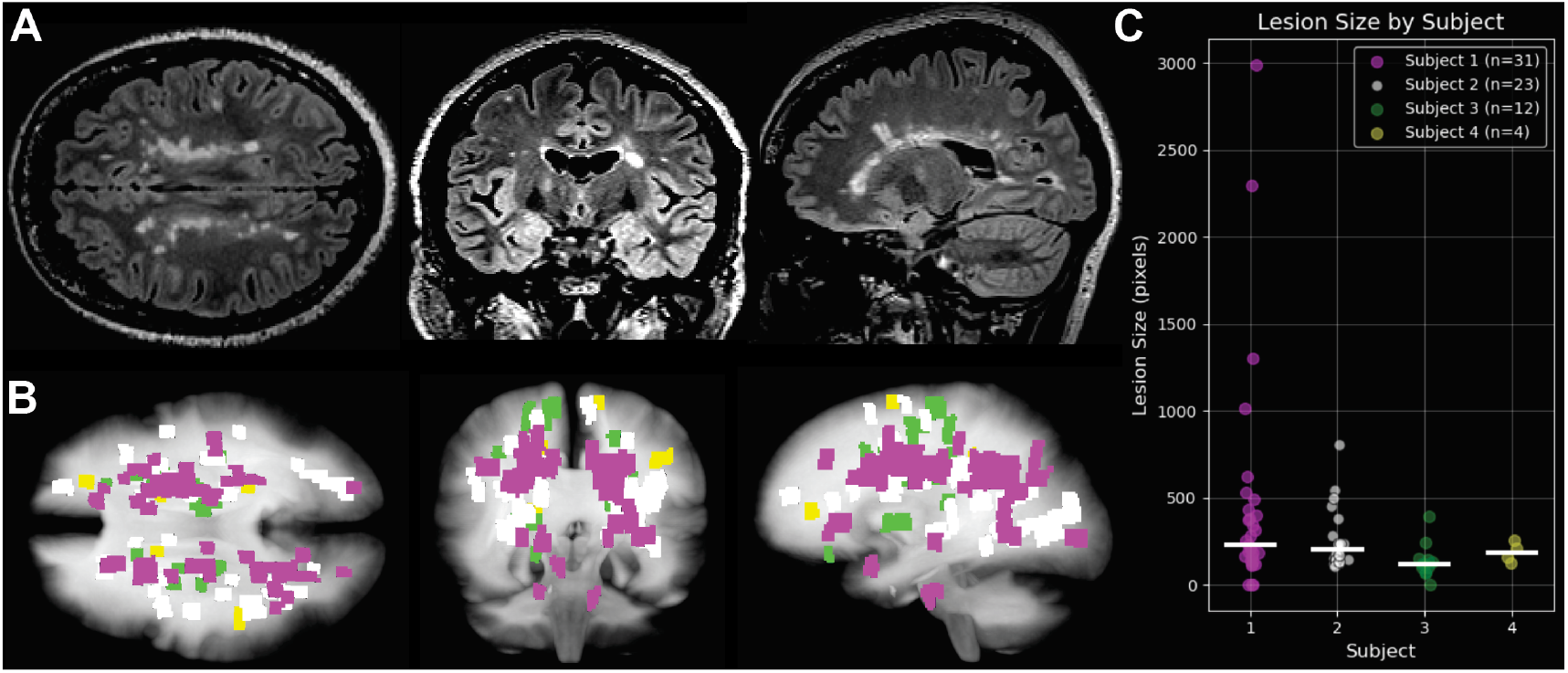
Anatomical MRI resolves MS lesions as hyperintense contrast. **A**: A representative T2-FLAIR MRI reconstruction of Subject 1. White matter appears hypointense compared to gray matter, with lesions presenting high positive contrast **B**: Rendering of MS lesions from 4 subjects manually segmented from MRI and registered to a reference atlas. **C**: Comparison of MS lesion sizes across 4 subjects (n= number of separable lesions)

### MRSI returns thousands of spectra that are low SNR and prone to artifacts

In a visual rendering of MRSI data, we examined a reference dataset, acquired without water suppression and summed all the spectral data into the equivalent of proton-weighted echo planar imaging (EPI) (**Figure 2A**). This showed a hypointense region around the sinuses and oral cavity, typical of susceptibility artifacts, and a posterior hyperintense region, typical of B1 field inhomogeneity

**Figure 2:**
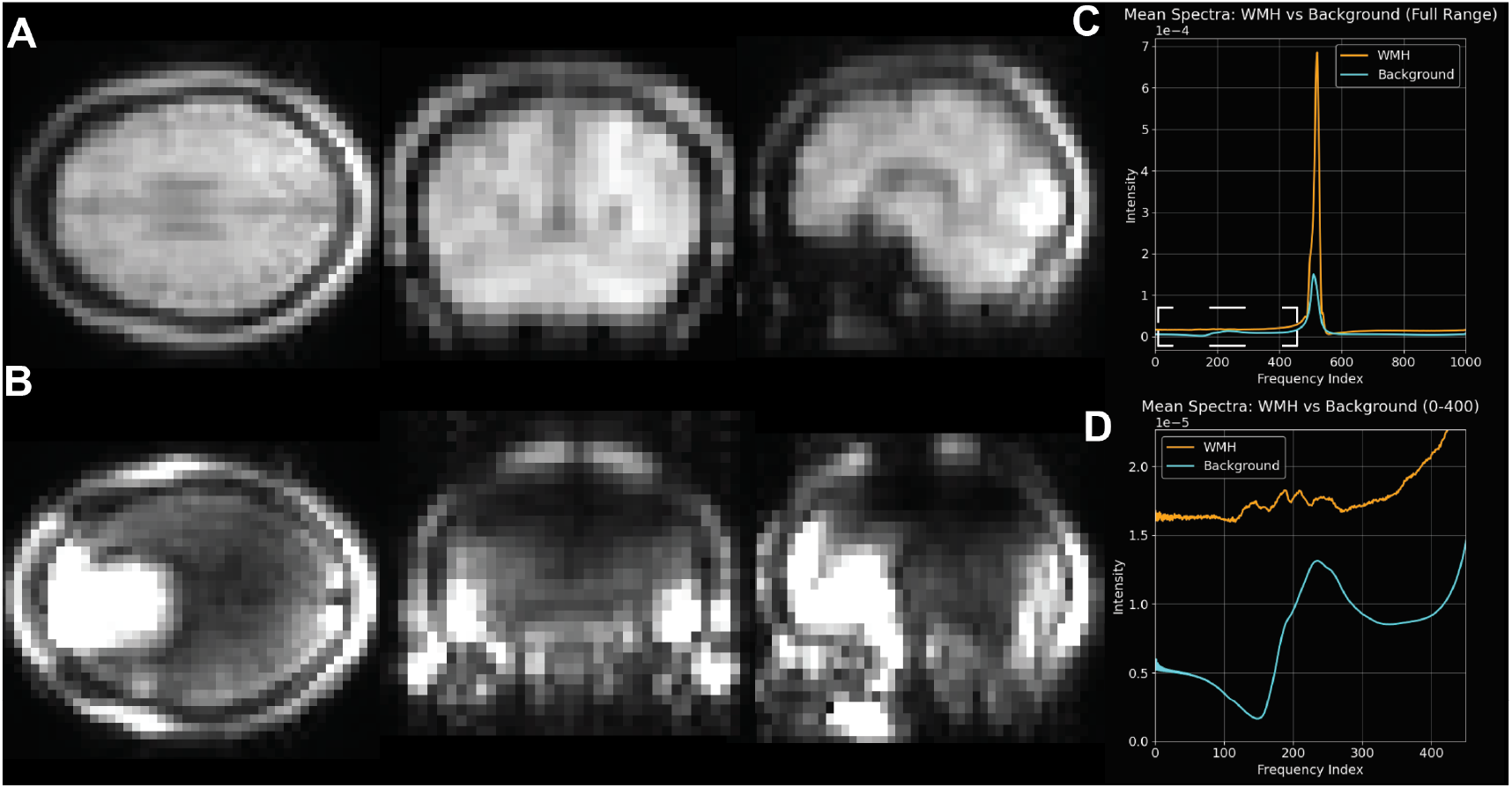
Spectral sums of MRSI reveal inhomogeneities and a dominant free water artifact. **A**: Spectral sum of reference MRSI data acquired without water suppression. **B**: Spectral sum of water suppressed MRSI acquisition. **C**: Mean spectra of all WMH regions vs Background non-lesioned brain. **E**: Upfield region (dashed line box) of WMH vs Background spectra.

EPSI methods partially suppresses water and lipid peaks but free water remains >100X more intense than any metabolite peak without additional suppression pulses (28). Reconstructing water suppressed MRSI and summing all spectral data as an image, demonstrated acquisition artifacts in the same regions as reference acquisition. In this case, both anterior and posterior brain regions appeared hyperintense, along with the ear canals (**Figure 2B**).

We then asked if MRSI spectra from WMH had some qualitative differences from the rest of brain tissues. After manual segmentation of whole brain and WMH regions from our anatomical images and transferring segmentation labels to our MRSI data, we plotted mean spectra of WMH regions vs the remaining brain. We did not exclude any brain regions based on the spectral sum reconstructions to avoid introducing bias beyond the initial labels. Comparing mean spectra from WMH vs general background brain tissue, the water peak from incomplete suppression dominated both labeled groups, appearing larger in the WMH labeled tissues (**Figure 2C**). Focusing on the upfield signal, some features were detectable, however averaged across so many spectra, chemical species could not be deconvolved and in the case of background brain, all chemical information blended into one spectral feature (**Figure 2D**). For WMH spectra, several peaks could be determined, however given the low spectral resolution, they could not be confidently associated with known metabolites.

### Imperfect water suppression is the dominant feature in MRSI data

We then asked if these MRSI spectra contained enough usable information in the available low-resolution features to separate without priors. We used PCA on minimally processed, water suppressed MRSI to reduce spectral data to a minimal set of vectors. 51% of the variance in the data could be attributed to the first 2 principal components (PC1,PC2). Representing all spectra along these axes however gave a singular cluster with a gaussian distribution (**Figure 3A**).

**Figure 3:**
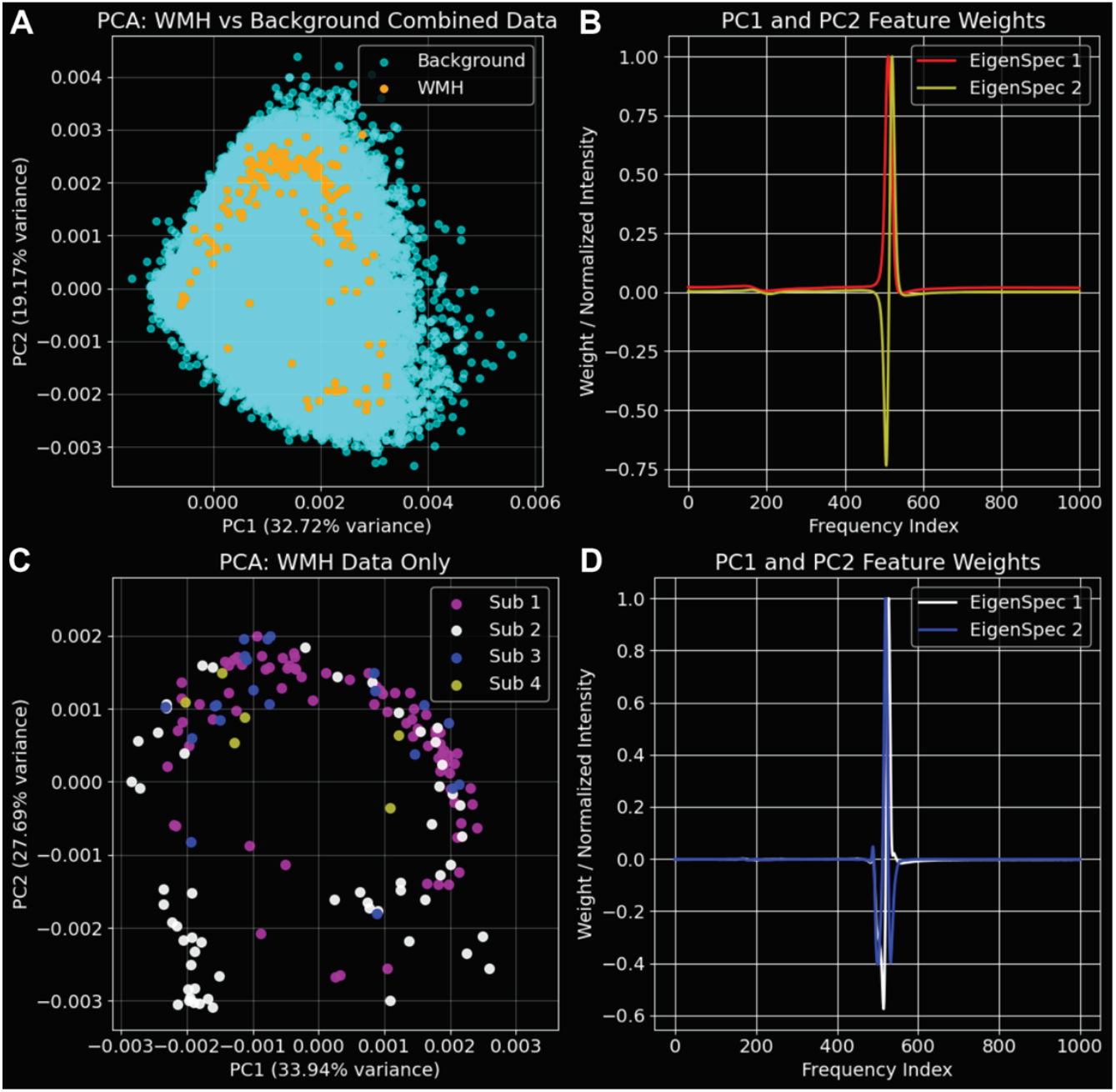
Principal component analysis of MRSI spectra. **A**: PCA of 163,000 brain spectra with labels. **B**: Single value decomposition of MRSI and reconstruction of the eigenspectra represented by PC1 and 2. **C**: PCA of 162 labeled WMH spectra. **D**: Eigenspectra represented by the first 2 principal components in C.

A pervasive issue in MRSI and NMR is the overwhelming free water signal, with any field inhomogeneity causing incomplete suppression of free water. To explore how prevalent the water signal was as a basis for separation in PCA, we used single value decomposition (SVD), the matrix decomposition used by PCA, to construct a plot of the feature weighting described by each component, or “eigenspectra” (**Figure 3B**). Plotting the eigenspectra of PC1 and PC2 showed the water peak as the main feature, indicating little other information contributed to separation of spectra along PC1 and PC2.

### Classification of WMH spectra is dominated by free water signal

MRI methods are already highly sensitive to WMH’s, and we asked if this prior knowledge would allow us differentiate between the lesions themselves. PCA on WMH voxels showed a similar distribution as the full brain PCA (**Figure 3C**). Plotting the eigenspectra of PC1 and PC2 again showed a dominant contribution by the free water signal, however the weighting was different from SVD of the full brain dataset (**Figure 3D**). While a simple PCA of WMHs were potentially more separable than all spectra combined in PCA space, it would be based on free water rather than metabolic features, and better resolved as MRI contrast.

### Contrastive PCA excludes the signal from free water

To leverage both our prior knowledge of which spectra came from WMHs and which spectral features were independent of the biology we were attempting to parse, we employed cPCA. Applying this method presumes that WMH salient features are convolved with a background of brain features that can be sampled independently and subtracted from the overall variance. This allowed us to ask which spectral elements varied between WMHs, while ignoring features that varied across the brain and between subjects.

Decomposing WMH spectra and plotting along 2 contrastive principal components (cPC1, cPC2) gave high overlap between subjects, with 3 distinct directions of variance (**Figure 4A**). K-means Clustering was sufficient to separate spectra (**Figure 4B**). Only one cluster was unique to a specific subject with otherwise high subject overlap (**Figure 4C**). Spectra from the subject with the least number of WMH voxels, representing a single lesion, fell into one cluster.

**Figure 4:**
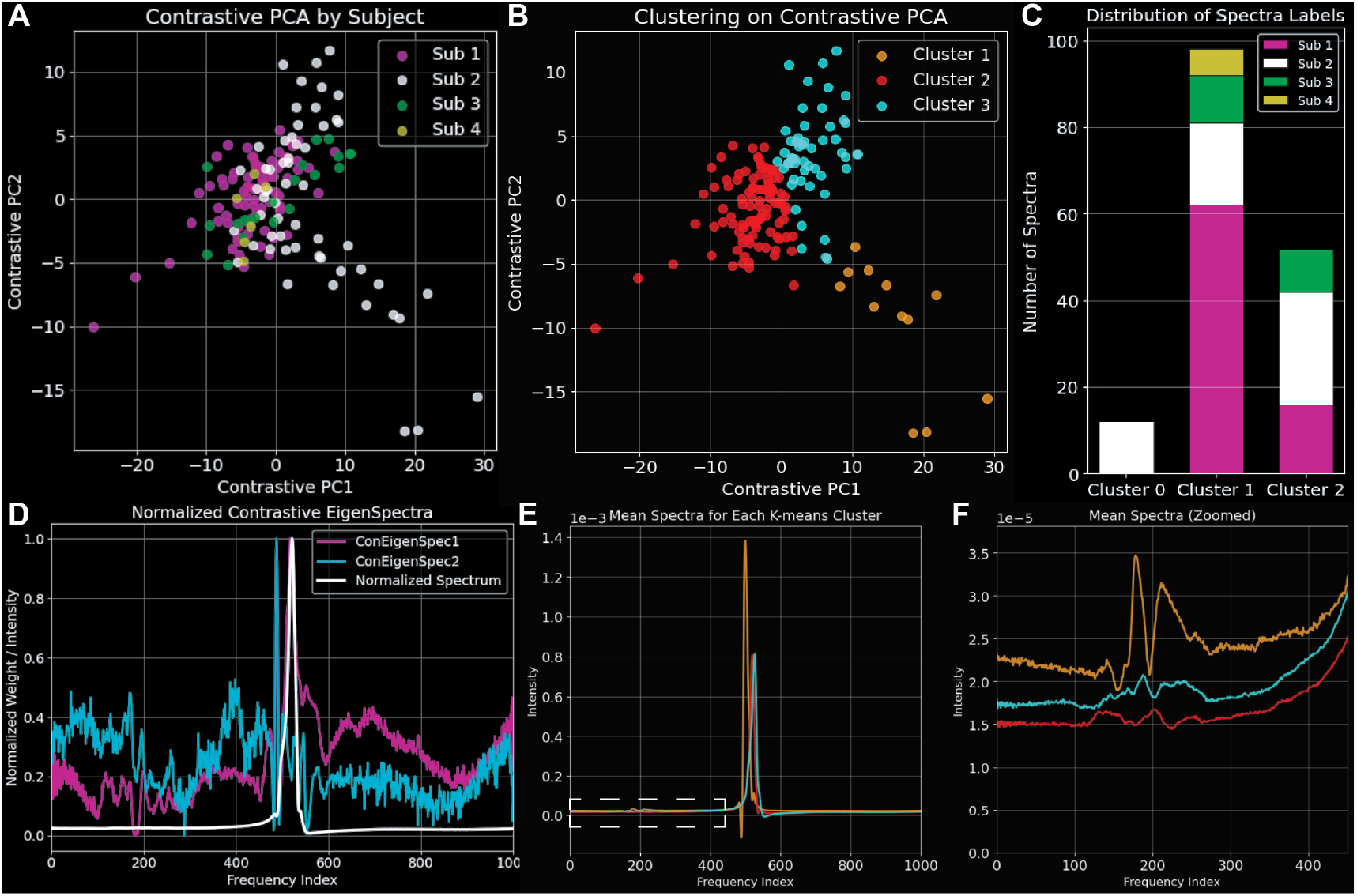
Contrastive PCA of WMH vs Background MRSI. **A:** Contrastive principal components of WMH spectra against background labeled by experimental subject. **B:** Clustering of cPCA data with labels. **C:** Distribution of clustered labels across subjects. **D:** Contrastive eigenspectra compared to global mean spectrum. **E:** Mean spectra of each cPCA cluster. **F:** Upfield region (dashed line box) comparison of mean cPCA clusters.

To explore which spectral features drove separation in cPCA, we reconstructed eigenspectral represented by cPCA1 and 2 (**Figure 4D**). Contrastive eigenspectra showed a balanced weighting across the whole spectra, however subregions of the water peak still contributed to each contrastive principal component. More surprisingly, far upfield and downfield regions were strongly weighted, despite having low variance in the actual spectra.

We then asked if clustering based on cPCA could yield visible features from whole spectra. Comparing mean spectra from each cluster still showed high water signal, with visible differences in the water peak between groups (**Figure 4E**). Focusing on the upfield region showed better resolved spectral features between clusters, with higher signal overall from Cluster 1 (**Figure 4F**).

### Lesions clustered by cPCA have distinct anatomical properties

To see if the separable differences across clusters were driven by anatomy, we applied clustering labels to anatomically segmented WMHs (**Figure 5A**). Comparing the spatial arrangement of the 3 separable clusters from background, there was no obvious gradient or clear association with artifactual regions identified by the spectral sum image. Cluster 1, found only in 1 subject, was the most posteriorly distributed group (**Figure 5B**), however Cluster 2 WMHs could be found even farther posterior. Cluster 2 overall appeared more distally distributed in every direction except ventrally and more anatomically separated from gray matter (**Figure 5C**). Cluster 3 was common to larger white matter tracts and deep brain regions, with significantly larger lesions than Cluster 2 and more lesions close to the center of the brain (**Figure 5D-F**).

**Figure 5:**
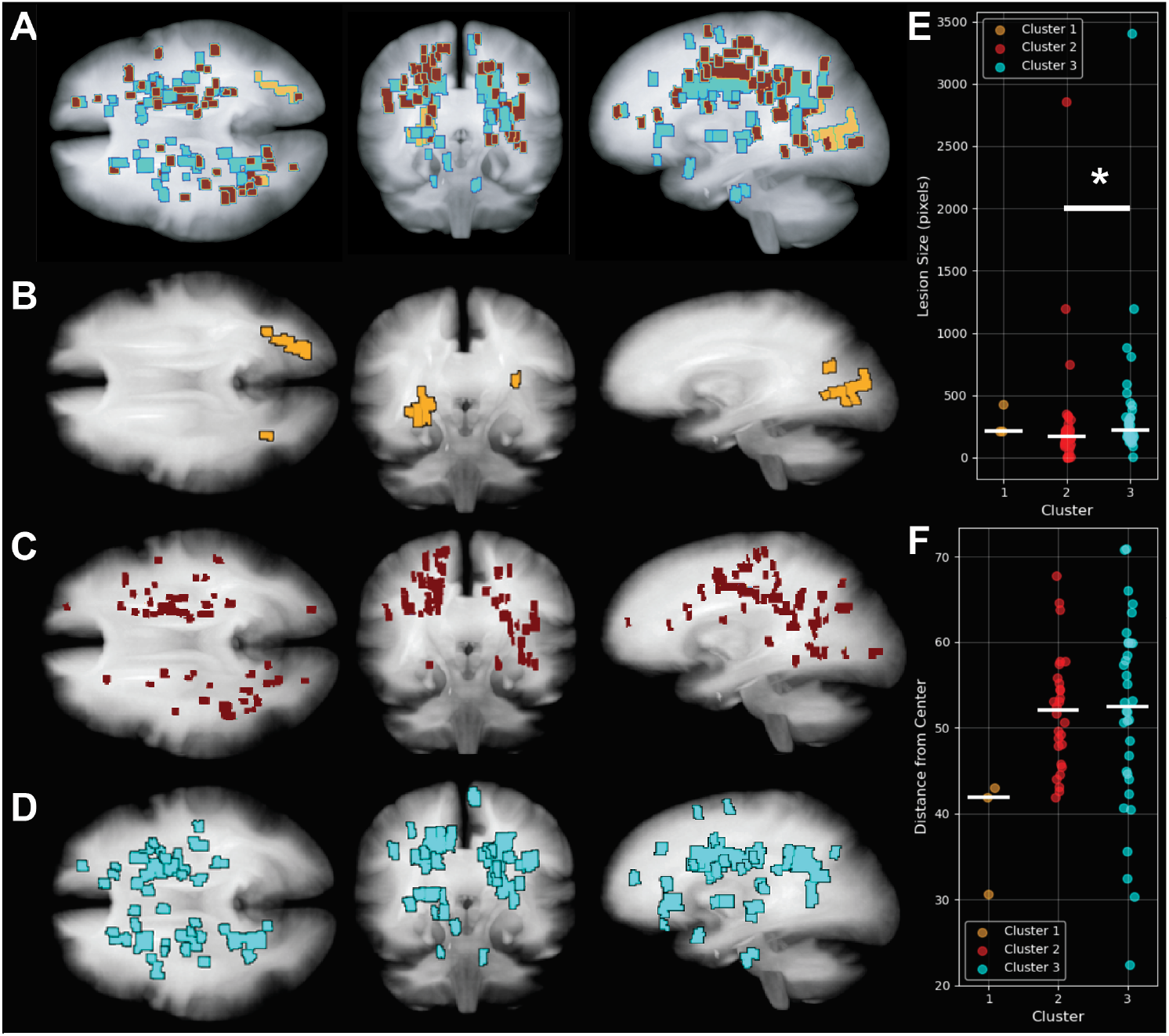
Projection of cPCA clusters into atlas space. **A:** Composite of all clusters in atlas space. **B:** Cluster 1 appears only in the subject with the highest lesion burden and exclusively in the posterior cortex. **C:** Cluster 2 appears in all subjects. **D:** Cluster 3 shows a more centralized distribution and is present in deep brain regions. **E:** Plot of lesion size based on cPCA

## DISCUSSION

MRSI offers an unprecedented scale of molecular information acquired noninvasively from human subjects. Here we explored the major impediments to effective analysis of MRSI data and present methods for coping with these challenges. Leveraging anatomical information for informed contrastive learning on MRSI data gives us the statistical power necessary to utilize machine learning approaches, while navigating artifact-prone data that is ambiguous to the human eye.

Based on contrastive features, we separated detectable lesions into 3 statistically significant clusters. Cluster 0 could only be found in the subject with the highest lesion burden, appearing only in posterior white matter as a large contiguous lesion. In contrast, cluster 1 lesions were as small as a single MRSI voxel and peripherally distributed. They were the only class of lesion to appear in Subject 4, who had the smallest burden of detectable lesions, suggesting they were lower severity than

In these data, areas prone to artifacts presented large free water signal that dominated unsupervised feature learning approaches. This was apparent in PCA-derived eigenspectra, which showed component weighting dominated by free water. One approach to filtering these data would be excluding the entire region around the water peak from analysis. In this analysis however, regions adjacent to free water were represented in cPCA-derived eigenspectra and therefore contributed to the first two contrastive principal components. Rather than setting arbitrary cutoffs, contrastive feature learning subtracts variance based exclusively on prevalence in background vs enrichment within samples, maximizing the information available based on the fewest assumptions. This adjunctive framework improves interpretability in artifact-prone regions and offers complementary mechanistic insight beyond vascular permeability alone.

While signal is molecular, we make no attempt to deconvolve specific metabolites from these data. It may well be possible to filter for metabolite specific signal, however the lack of any clear peaks would leave high uncertainty in the analysis (19). The clear study design allows for a testable hypothesis regarding lesion-specific differences, but this same approach would not be applicable to questions of brain-wide inflammation (29). Here, non-lesion brain tissue serves as a crucial internal normalization factor and filter for artifacts. We do not yet know if contrastive learning is compatible with inter-subject comparisons. Appropriate datasets for this work are being actively pursued by the metabolomics community (30).

While clinical MRI is effective at detecting the physical signs of inflammation through dynamic contrast and diffusion-weighted imaging, without the benefit of molecular markers, contrast changes are ambiguous. With the right framework, MRSI presents a molecularly resolved assay of neuroimmune responses, reducing misdiagnosis and guiding early treatment of neuroinflammatory states. MRSI requires high field (3T) systems for adequate SNR and spectral resolution, but MRI is on a steady trend towards higher field strengths. There is a growing availability of ultra-high field (7T+) human research scanners, with some as powerful as 20T (31, 32). These systems have demonstrated value for fMRI studies of hemodynamic responses, but we have only just begun to tap their potential for metabolic imaging.

## Data Availability

All inquiries regarding data access should be directed to the Mayo Clinic Institutional Review Board (IRB).

## DATA AND CODE AVAILABILITY

The data that support the findings of this study are subject to institutional and ethical restrictions. All inquiries regarding data access should be directed to the Mayo Clinic Institutional Review Board (IRB). Deidentified spectra and all code used for registration and analysis are available on Zenodo. Reviewer Link: https://zenodo.org/uploads/18306067?token=eyJhbGciOiJIUzUxMiIsImlhdCI6MTc2ODg2MTU2MiwiZXhwIjoxNzk4NzYxNTk5fQ.eyJpZCI6IjcwMmIwYjBmLWU4NTMtNGU0Ni05NzUxLTUwMDQ1YjBmYjBmNSIsImRhdGEiOnt9LCJyYW5kb20iOiI1YjEyYjJmZDJkZmI4ZDhhNzFkMjdiMzdjY2RjOTI0ZCJ9.Cxa_v97T1_8wqtmfoX0HCSTFyI_jMzvxC5r6xQjbHDwamEXY5ZCCDzb_TjjeIV7jPsdUM2op8MtQYmskPZqPIA

Dataset DOI for public release upon acceptance: 10.5281/zenodo.18306067

All analysis scripts are available on GitHub https://github.com/BioProteanLabs/MRSI_cPCA_MayoMS

## Notes

Publication statement: Conflicts of interest: The authors have no competing financial interests.

### Competing Interest Statement

The authors have declared no competing interest.

### Funding Statement

This studey was funded by NIH Office of the Director DP2MH136493

### Author Declarations

The research performed and data generated for this study were approved by the Mayo Clinic Institutional Review Board and all data remains subject to institutional and ethical restrictions of Mayo Clinic

